# MRI measures of cortical microstructural damage associate with tau accumulation in presymptomatic Alzheimer’s disease

**DOI:** 10.64898/2025.12.15.25340517

**Authors:** Philip SJ Weston, William Coath, Thomas Brown, Catherine J Scott, Ian B Malone, Erik Årstad, Ramla Awais, Kerstin Sander, David L Thomas, John Dickson, Michael Schöll, Marcus Richards, Frederik Barkhof, Nick C Fox, H Zhang, David M Cash, Jonathan M Schott

## Abstract

**Objectives:** Amyloid lowering therapies for Alzheimer’s disease are most effective in those without significant neocortical tau spread. Currently, the only method for anatomically staging tau is through PET scanning, which is largely unavailable in clinical settings. We assessed whether advanced MRI biomarkers of cortical microstructure may provide a more accessible option for assessing early neocortical tau accumulation.

**Methods:** 217 asymptomatic individuals underwent amyloid-β PET, tau PET and multi-shell diffusion 3T MRI. Neurite orientation dispersion and density imaging (NODDI) metrics including orientation dispersion index (ODI; a proxy measure of dendritic complexity), and free water fraction (FWF) were calculated. Analysis focused on a meta-temporal cortical composite region, the first site of neocortical tau beyond the medial temporal lobe (MTL).

**Results:** Meta-temporal tau burden was associated with meta-temporal ODI (Pearson’s r=-0.24, p=0.001), with a weaker association also seen with meta-temporal FWF (r=-0.14, p=0.07). Within the amyloid-β positive group, ODI discriminated between those with and without meta-temporal tau positivity (area under the curve=0.86).

**Discussion:** NODDI-derived ODI is associated with tau PET signal, even in preclinical disease and may discriminate individuals with-and without tau beyond the MTL. NODDI represents a potential alternative imaging tool for staging tau spread, aiding future treatment stratification and delivery.

## Introduction

As Alzheimer’s disease (AD) enters an era of disease modifying treatments,^1 2^ early diagnosis and disease staging are crucial, with therapies likely to be most effective if given prior to the development of significant cognitive decline.^3^ While evidence of amyloid-β (Aβ) deposition is a core eligibility requirement for anti-amyloid immunotherapy, mapping the extent of hyperphosphorylated tau spread, in-line with recently updated staging criteria,^4^ provides additional stratification to determine who is most likely to benefit.^2^ Tau deposition begins in the medial temporal lobe (MTL) before spreading into the neocortex,^5^ with those without significant neocortical tau having better treatment outcomes.

Currently, the only method available for anatomical tau staging is positron emission tomography (PET),^6^ which is expensive, involves radiation, and has limited clinical availability, precluding use at scale. Alternative imaging approaches able to provide a proxy measure of cortical tau are therefore required.

Neurite orientation dispersion and density imaging (NODDI), an advanced form of diffusion MRI, has shown potential for estimating cortical tau spread. NODDI enables investigation of disease-related processes, including estimation of dendritic structural complexity through quantification of the orientation dispersion index (ODI).^7^ We have previously shown in individuals with AD dementia that ODI is closely associated with regional tau deposition.^8^ Others have highlighted the association between tau deposition and a second NODDI measure, the free-water fraction (FWF).^9^

In the current study, we investigate the association between tau PET binding and both ODI and FWF during the presymptomatic stage of AD, with a particular focus on how well NODDI measures discriminate between those with and without early neocortical tau involvement

## Methods

### Participants and procedures

We included 217 asymptomatic individuals from the MRC National Survey of Health and Development (NSHD; the British 1946 Birth Cohort).

All participants underwent combined PET/MR with Aβ-PET at mean age 73.0 (standard deviation 0.7) and tau-PET at age 77.2 (0.9). All images were acquired on the same 3T PET-MRI scanner (Siemens Healthcare). The Clinical Dementia Rating Scale (CDR) assessed cognitive and functional status.

[18F]florbetapir Aβ-PET used acquisition and processing methods as described previously.^10^ A 12 Centiloid cut-point for amyloid status was determined using a mixture modelling approach based on the 99th percentile of the lower (amyloid-negative) Gaussian.

[18F]MK-6240 tau PET standard uptake value ratios (SUVRs) were calculated using an inferior cerebellar reference region, with tau positivity defined using the same mixture model approach (SUVR>1.255).

MRI was acquired concurrently during tau PET, including 3D T_1_-weighted MPRAGE (TI/TR=870/2000ms, voxel size 1.1mm^3^) and multi-shell dMRI (TR=8000ms, TE=103ms, voxel size 2.5mm^3^) with b-values of 2,000 (64 directions); 700 (32 directions) and 0s/mm^2^ (12 repetitions). Cortical grey matter parcellation of T_1_-weighted images was performed using GIFv3,^11^ with subsequent resampling into amyloid PET, tau PET and dMRI space. The NODDI model provided both ODI and FWF.

### Statistical analyses

In line with our study aim, analysis was restricted to a single meta-temporal composite region, representing the first site of neocortical tau beyond the MTL. We used Pearson correlation coefficients to quantify associations between tau PET SUVR and ODI and FWF. Where associations were found, linear regression assessed whether they remained after adjustment for 1) amyloid status, 2) cortical volume, and 3) the other NODDI measure (i.e. for ODI, adjustment for FWF and vice-versa). Receiver operating characteristic (ROC) curves were generated using logistic regression to quantify the ability of ODI and FWF in amyloid-positive individuals (those in whom tau staging would be required) to discriminate between those with and without meta-temporal tau deposition.

## Results

### Participant demographics, based on Aβ and tau status, are outlined in Table 1

Across the entire cohort, meta-temporal tau was significantly associated with meta-temporal ODI (Pearson’s r=-0.24, p=0.001), and weakly with meta-temporal FWF (r=-0.14, p=0.07) (Figure1). The association between ODI and tau persisted after adjusting for amyloid (p=0.004), regional volume (p=0.03), and FWF (p=0.005). Only including Aβ positive individuals, correlations were stronger: ODI and tau, r=-0.40, p=0.001; FWF and tau r=0.30, p=0.017.

**Figure 1.**
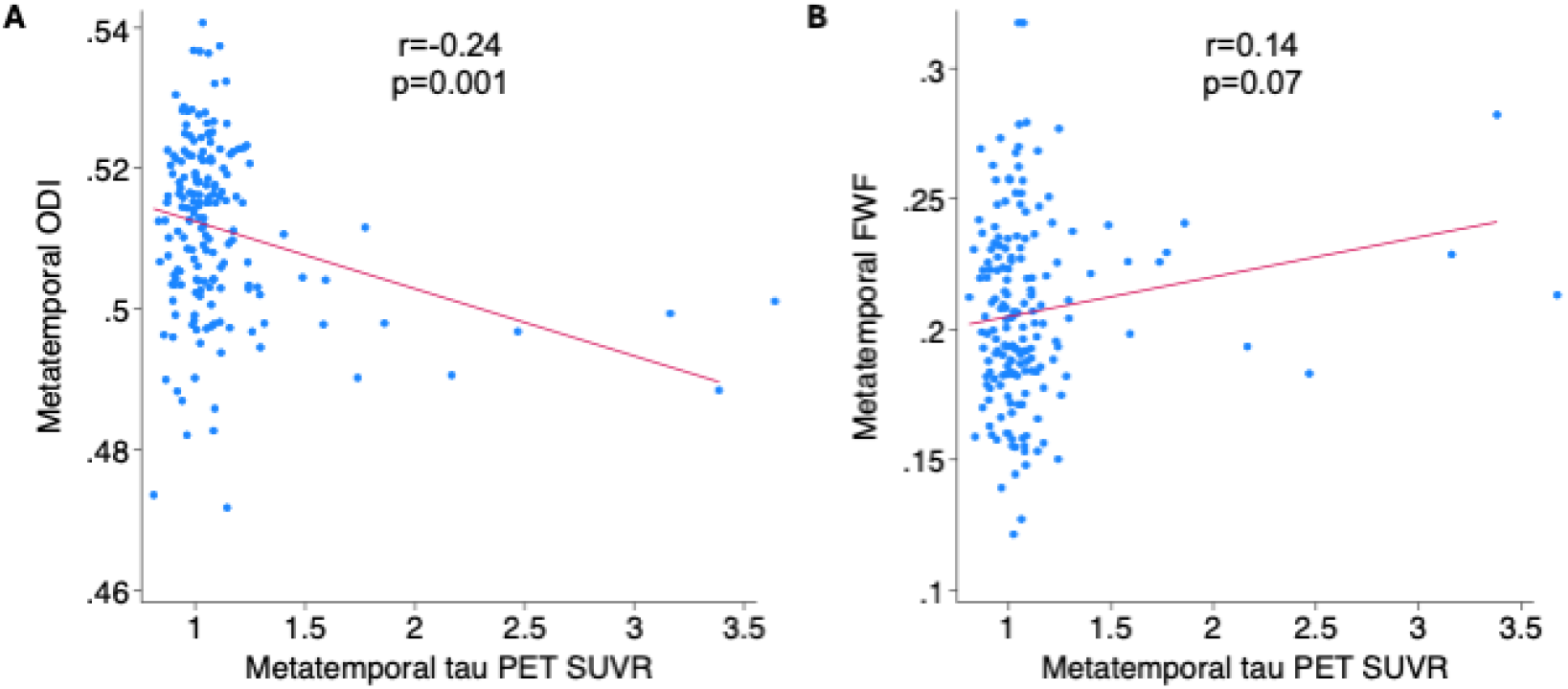
Association between regional tau PET and NODDI measures. Scatter plots show the association, in the meta-temporal region, between tau PET signal and A) ODI, and B) FWF, across the entire cohort. Pearson’s r and corresponding p-values are shown.

**Table 1.** Demographics of the cohort, split based on amyloid a tau positive/negative status. A= amyloid-β, T = tau. CDR = Clinical Dementia Rating scale.

|  | A- / T- | A+ / T- | A+ / T+ |
| --- | --- | --- | --- |
| N | 141 | 42 | 32 |
| Age, mean (SD) | 77.5 (0.9) | 76.6 (0.8) | 76.9 (0.7) |
| Sex (m/f) | 75/64 | 21/22 | 19/14 |
| CDR, median (interquartile range) | 0 (0-0) | 0 (0-0) | 0 (0-0) |
| Mean global amyloid (centiloids), mean (SD) | -1.5 (5.7) | 38.5 (24.9) | 63.3 (32.4) |
| Meta-temporal tau (SUVR) | 1.00 (0.08) | 1.03 (0.09) | 1.47 (0.45) |
| Meta-temporal ODI | 0.513 (0.012) | 0.515 (0.012) | 0.504 (0.012) |
| Mean meta-temporal FWF | 0.206 (0.035) | 0.200 (0.036) | 0.216 (0.030) |

Within the Aβ positive group, ODI was able to discriminate between those with/without meta-temporal tau positivity (area under the curve [AUC]=0.86 [95% confidence interval 0.77-0.95], sensitivity=0.87, specificity=0.78) but this was not the case for FWF (AUC=0.62 [0.47-0.77], sensitivity=0.60, specificity=0.63) (Figure 2).

**Figure 2.**
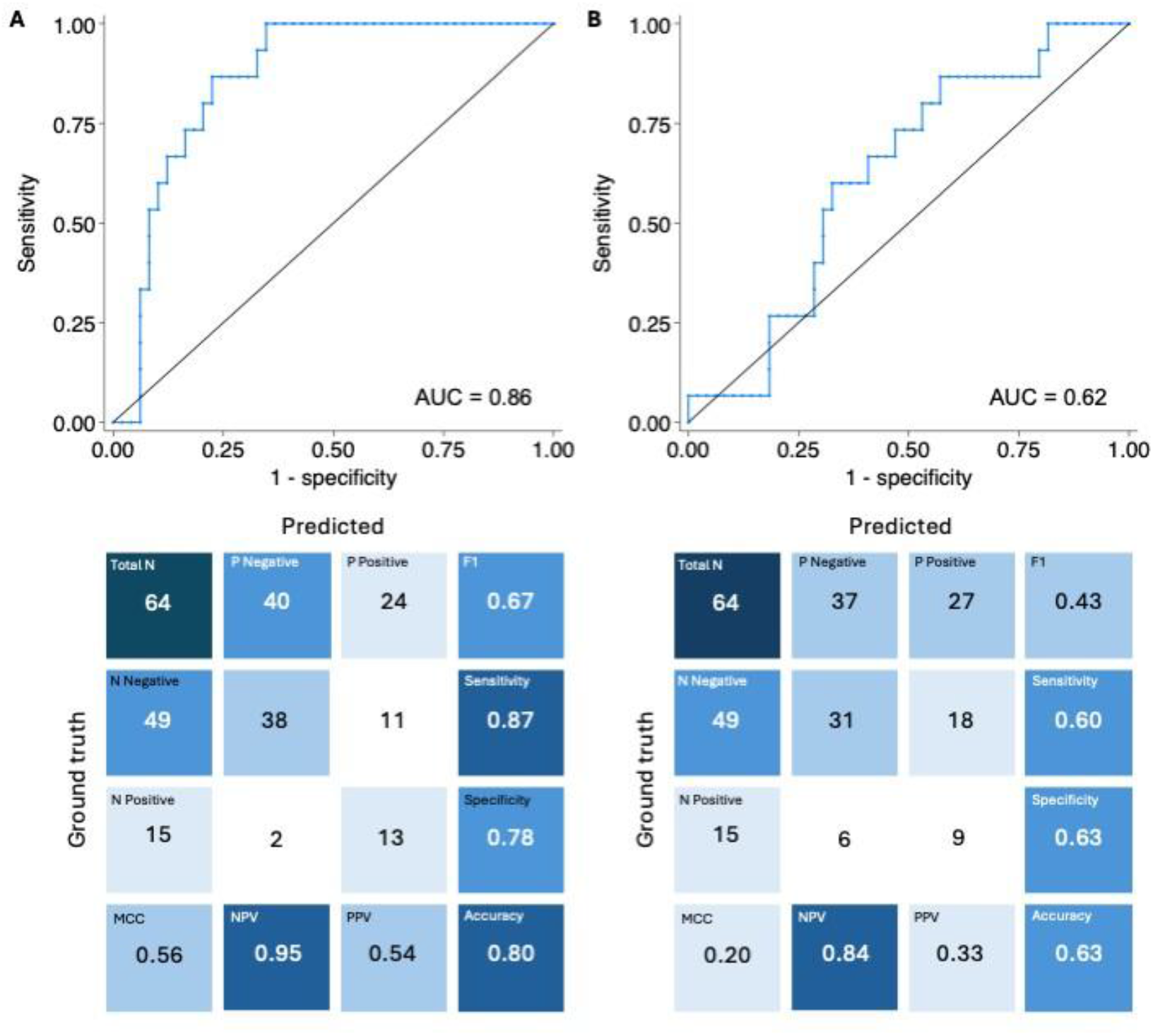
Ability to discriminate those who do and do not have significant tau spread. Receiver operating characteristic curves and associated confusion matrices for both ODI and FWF, showing the ability to discriminate between amyloid positive people with and without tau spread beyond the medial temporal lobe. AUC = area under the curve. MCC = Matthews Correlation Coefficient. NPV = negative predictive value. PPV = positive predictive value.

## Discussion

In a cohort of non-demented elderly individuals, we found that NODDI, and in particular regional ODI, is associated with tau PET binding in temporal neocortex, a brain region that is usually the first site of tau spread beyond the MTL. Moreover, in those who would be potentially eligible for anti-amyloid therapies (i.e. Aβ positive), ODI provides good discrimination between those with and those without tau beyond the MTL.

Previous AD studies found significant associations between tau PET and both ODI and FWF, with both microstructural measures more closely reflecting underlying tau than macrostructural thickness/volume measures.^8 9^ However, these studies were performed in relatively small samples containing individuals who had already developed progressive cognitive decline. The current study builds on previous findings by investigating ODI and FWF concurrently, in a larger cohort of individuals at an earlier disease stage.

The association between tau and ODI was independent of regional brain volume and regional free water, consistent with ODI – a marker of dendritic branching – providing more structurally specific information relating to tau deposition. This finding is supported by a non-human study that found focal tau deposition to cause loss of dendritic spines and reduced arborization.^12^ While ODI appeared to closely reflect regional tau deposition, the association between tau and FWF was weaker. FWF, which reflects the proportion of a voxel occupied by free water (the inverse of the proportion occupied by neural tissue), is not specific to any particular component of the degenerative process, likely explaining its lower sensitivity and specificity to early tau accumulation.

Anatomically staging tau, particularly prior to significant clinical decline, may support future delivery of disease modifying treatment at large scale. It has been recently shown that the anatomical spread of tau is driven by different processes than overall tau load, with the two measures (spread versus load) not being directly interchangeable.^13^ The necessity to identify *where* rather than simply *if* tau is present underpins the need for cost-effective and scalable alternative imaging methods to tau PET, with NODDI offering a potential solution. NODDI can already be acquired on a standard 3T MRI scanner, with recent technical advances enabling 1) acquisition over shorter time periods (∼5 minutes),^14^ and 2) on-scanner automated generation of NODDI maps^15^ making it a feasible addition to a standard clinical protocol.

Our study has some limitations. There was a gap of approximately four years between Aβ PET and tau PET acquisitions raising the possibility that some participants may have developed amyloid positivity between the two scans. While the overall cohort size is large, only a minority of participants were tau positive, reflecting the overall early clinical stage of the cohort. However, the fact that NODDI metrics showed close association with tau even in the relative absence of widespread pathology further emphasises its potential value.

In summary, we show that cortical NODDI, and particularly ODI, is associated with regional tau PET, even at the preclinical disease stage. ODI can discriminate between Aβ positive people with and without tau spread beyond the MTL. NODDI is a potential scalable, non-invasive imaging tool for staging of tau spread, aiding future treatment stratification and delivery.

## Data Availability

Data from the NSHD are curated and stored by the MRC Unit for Lifelong Health and Ageing at UCL. Anonymized data will be shared by request from bona fide investigators (https://skylark.ucl.ac.uk/ NSHD).

